# Moderate to severe negative symptoms predict low risk of symptoms worsening in schizophrenia patients in CATIE

**DOI:** 10.64898/2026.02.07.26345806

**Authors:** Helene Speyer, Jonathan Rabinowitz, Remy Luthringer, Bogdan Ionel Tamba, Michael Davidson

**Affiliations:** Grigore T. Popa University of Medicine and Pharmacy, Ia□i, Romania; Mental Health Services in the Capital Region of Denmark; Bar-Ilan University, Israel; Minerva Neurosciences, USA

**Keywords:** Schizophrenia, Negative symptoms, Symptom exacerbation, Relapse prediction

## Abstract

Understanding factors that predict the course of schizophrenia remains essential for improving long-term clinical management. Rate and severity of symptom exacerbations vary widely across individuals, and although prior studies have examined potential predictors, findings have been inconsistent and often limited by small samples, infrequent assessments, and non-standardized measures. Using data from phase 1 of the Clinical Antipsychotic Trials of Intervention Effectiveness (CATIE), which includes a large cohort with monthly standardized evaluations, this study investigated whether baseline negative symptom severity predicts risk of symptom exacerbation over time. Participants were 1139 adults aged 18–65 years meeting DSM-IV criteria for schizophrenia. Symptoms worsening or exacerbation was defined as a ≥12-point increase from baseline on the PANSS total score. Cox regression survival models examined the association between baseline PANSS negative symptom tertiles and time to exacerbation, adjusting for age, sex, PANSS positive and general psychopathology subscales, and CGI-Severity. Overall, 25.5% of participants experienced exacerbation over a 18-month period of follow-up. Survival curves demonstrated significant separation across negative symptom tertiles (p=0.047), with higher baseline negative symptoms associated with longer time to exacerbation. Compared with the lowest tertile, medium and high negative symptom groups showed reduced exacerbation risk (HR=0.73 and HR=0.69, respectively; both p=0.03). Findings indicate that greater baseline negative symptom severity is associated with a lower likelihood of short-term symptom worsening, suggesting a relatively stable illness course among individuals with more severe negative symptoms. These results have implications for prognosis and treatment planning, while underscoring the persistent functional burden imposed by negative symptoms despite lower exacerbation risk.

## 1. Introduction

The natural course of individuals defined by DSM as suffering from schizophrenia consists of psychotic symptoms worsening and ameliorations defined as exacerbation and for many, persistent negative symptoms, cognitive impairment and social dysfunction. The intervals between exacerbations vary between patients. To provide the best clinical care clinicians and researchers are investigating phenomenological and biological indicators which might predict medium- and long-term course.

While some studies have found no predictive value of clinical variables (Hansen et al., 2024), others indicated that substance abuse, smoking status, severity of positive symptoms, large number of hospitalizations, high levels of depressive and negative symptoms, and low global functioning might offer a limited degree of predictive value (Barbosa et al., 2025; van Dee et al., 2023; Wunderink et al., 2020).

A recent report of 483 schizophrenia patients with moderate to severe negative symptoms, indicated that 90% maintained symptoms stability and avoided hospitalization for the next 9-12 months (Rabinowitz et al., 2023). To elaborate and confirm this finding in an independent sample, the analysis presented here utilized the CATIE (Swartz et al., 2008) dataset to examine rates of exacerbation by degree of negative symptoms.

Studies exploring predictors of symptom exacerbation have several limitations, including non-standardized measurements in naturalistic cohorts, long intervals between assessments that fail to capture what occurs in the interim (Lieberman and Stroup, 2011), and small sample sizes that increase the risk of spurious findings (Siafis et al., 2024). The CATIE (Clinical Antipsychotic Trials of Intervention Effectiveness) dataset addresses these limitations by providing a large sample with standardized monthly follow-ups. By examining the predictive role of negative symptoms in exacerbation, this study seeks to clarify their contribution to illness trajectory.

## 2. Methods

This is a longitudinal survival analysis of phase 1 of data from CATIE. Participants were adults aged 18–65 years meeting DSM-IV criteria for schizophrenia. Participants missing baseline negative symptom data or not having at least one follow-up assessment were excluded. The final analytic sample included 1,139 individuals. As proposed by Siafis et al (2024) in a major data informed expert consensus study, exacerbation or relapse was defined as a 12-point or greater increase from baseline on PANSS (Positive and Negative Syndrome Scale) total.

Data were analyzed using Cox regression survival analysis to examine how multiple factors together influence the time until an event (exacerbation) occurs, without assuming any specific pattern for how risk changes over time. Participants were categorized into tertiles based on their negative symptom level at baseline. Categories were used as independent variable using simple parameter coding, with the low negative symptom group serving as the reference category to which the medium and high negative symptoms groups were compared. This approach enabled estimation of hazard ratios for relapse for each higher severity group relative to the lowest group. Additional variables entered in the Cox regression were baseline PANSS Positive subscale, PANSS General psychopathology subscale, and Clinical Global Impression of illness severity (CGI-S), allowing estimation of the independent contribution of negative symptoms to relapse risk beyond other symptom domains. All models adjusted for age and sex. All analyses were conducted using SPSS (version 31).

## 3. Results

Mean (SD) PANSS total score was 75.3 (17.2), for positive subscale score was 18.2 (5.5), for negative subscale was 20.3 (6.4) and for general psychopathology was 36.8 (9.1). Approximately 25.5% (n=291) of participants experienced symptom exacerbation over a period of 18 months follow-up. Table 1 presents the background characteristics by negative symptom tertile group.

**Table 1.** Background characteristics of PANSS negative symptom tertile groups.

| Negative symptoms subscale tertile group | Sex | Age | PANSS positive | PANSS general psychopathology | CGI-S |
| --- | --- | --- | --- | --- | --- |
| Less than 17 (n=330) | Males 68.2% (n=225)<br>Females 31.8% (n=105) | 41.3 | 16.26 (5.5) | 30.5 (7.3) | 3.55 (.99) |
| 17 to 22 (n=419) | Males 75.9% (n=318)<br>Females 24.1% (n=101) | 40.8 | 18.4 (5.2) | 36.9 (7.9) | 3.94 (.84) |
| More than 22 (n=389) | Males 77.6% (n=302)<br>Females 22.4% (n=87) | 40.2 | 19.6 (5.3) | 42.0 (8.3) | 4.30 (.86) |
| P= | 0.01 | 0.39 | <0.001, d=0.51 | <0.001, d=1.1 | <0.001, d=0.67 |

Estimated survival curves as shown in Figure 1 demonstrated clear separation between the three baseline negative symptom groups, with the less severe negative symptom group showing the fastest decline in survival probability over time, and the most severe negative symptom group showing the most prolonged survival without exacerbation or study discontinuation (p=0.047). As compared to the lowest score negative symptom group, each of the two subsequent negative symptom groups had a lower rate of exacerbation (Exp(B)=.73, p=.03) and (Exp(B)=.69, p=.03). Thus, compared to those with negative symptom total at baseline of less than 17, those with scores 17 to 22 had a *reduced* odds of relapse by .73 and those with negative symptoms greater than 22 had a .69 *reduced odds* as compared to those less than 17.

**Figure 1.**
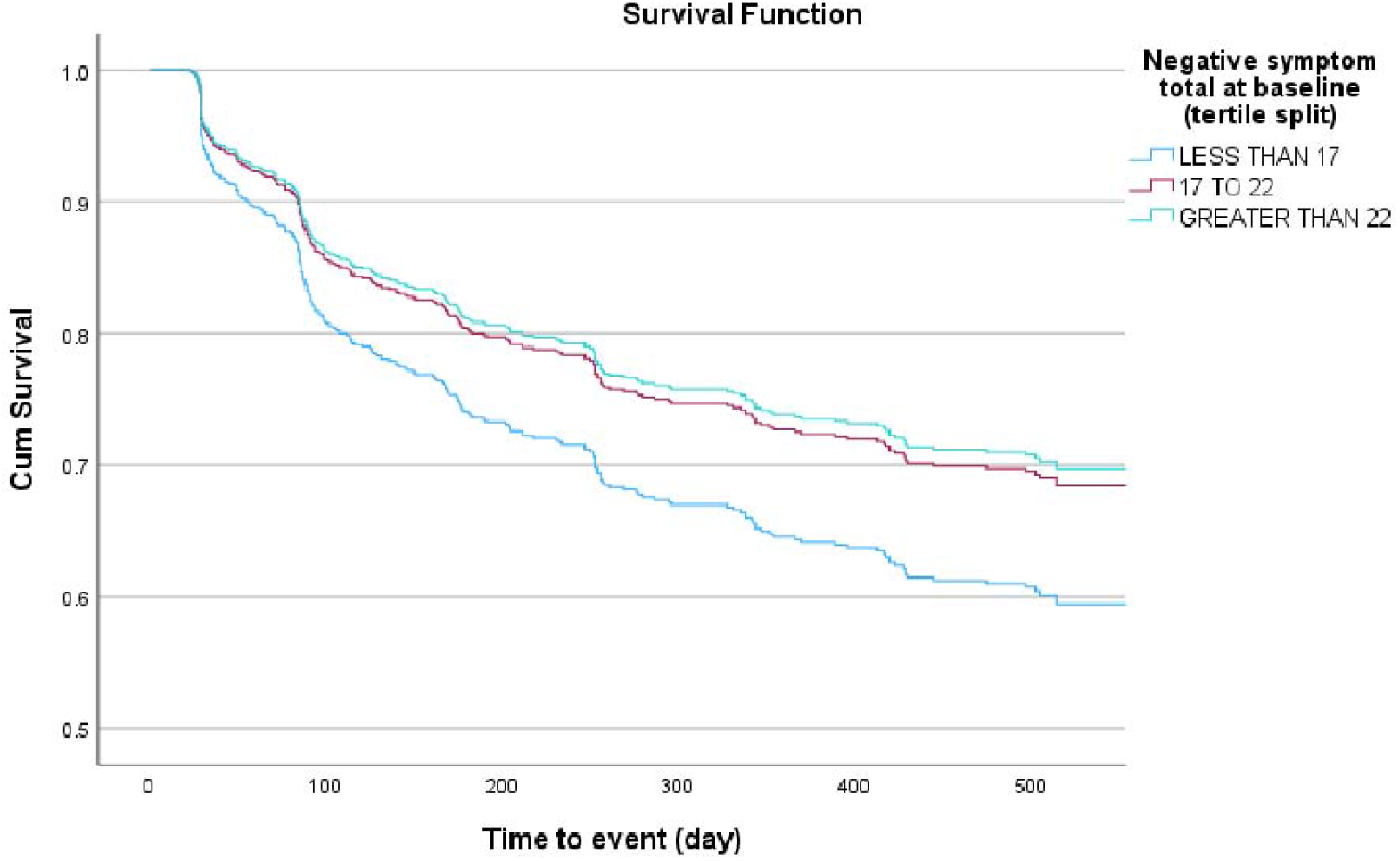
Cox regression survival curves of relapse-free survival by negative symptom severity. PANSS relapse criteria

## 4. Discussion

This study demonstrates that greater baseline negative symptom severity is associated with a lower risk of symptom exacerbation over time. These findings suggest that negative symptoms may identify a subgroup of individuals with schizophrenia characterized by relative stability.

Our results differ from prior studies reporting increased relapse risk among individuals with prominent negative symptoms (van Dee et al., 2023; Wunderink et al., 2020). This discrepancy may be explained by differences in relapse definitions, illness stage, and follow-up density. Whereas earlier studies often defined relapse by hospitalization, or studied first-episode cohorts, the present analysis examined symptom-based exacerbation in a chronic, well-characterized sample with frequent assessments.

The results reported here are consistent with those reported in individuals with moderate/severe negative symptoms treated for a similar period with Roluperidone, a compound with antagonist properties for 5-HT2A, sigma2, and α1A-adrenergic receptors but very limited Dopamine receptor blocking activity (Rabinowitz et al., 2023).

In another secondary analysis of the CATIE trial (Rabinowitz et al., 2012), negative symptoms were a significantly stronger predictor of functional outcomes than positive, disorganized, or depressive symptoms. The findings suggest that improvement in negative symptoms may have a distinctive and independent impact on functional outcomes in schizophrenia, supporting the importance of specifically targeting negative symptoms in treatment and in future research.

Several mechanisms may account for this pattern. Individuals with moderate to severe negative symptoms may exhibit lower behavioral reactivity to environmental stressors, limiting the emergence of acute psychotic exacerbations. Alternatively, negative symptoms may reflect a distinct biological phenotype with greater cognitive impairment and functional decline (Rabinowitz et al., 2012) but reduced episodic symptom fluctuation.

Clinically, these findings have implications for prognosis and treatment planning. Identifying patients with a lower risk of exacerbation could inform decisions about antipsychotic medication tapering, monitoring intensity, and psychosocial intervention prioritization. However, negative symptoms remain a major driver of disability, underscoring that lower relapse risk does not equate to better overall outcomes, highlighting the need for negative symptom treatments (Mittal et al., 2026).

Limitations to this study include the observational nature of the analysis, potential regression-to-the-mean effects for the portion of the variance in exacerbation due to negative symptom level, which we tried to control for by including CGI-S, and the inability to distinguish exacerbation occurring on versus off medication. Future studies should integrate symptom trajectories with medication exposure, biomarkers, and functional outcomes to refine schizophrenia phenotyping.

Our analysis of CATIE, suggests that higher baseline negative symptom severity predicts a reduced risk of psychotic relapse in schizophrenia, suggesting the existence of a clinical phenotype marked by symptom stability rather than recurrent exacerbation. These findings challenge prevailing assumptions and highlight the importance of symptom-based stratification in relapse prevention research.

## Data Availability

All data processed are available upon request to NIH https://nda.nih.gov/

https://www.nimh.nih.gov/funding/clinical-research/practical/catie

